# Implementing Inclusive and Community-Driven AI for Health programs: Lessons from Frontier Technology Pilots in Developing Contexts

**DOI:** 10.64898/2025.12.16.25342439

**Authors:** Christopher Moore, Julius Mugwagwa, Ian Vickers

## Abstract

The use of Artificial Intelligence (AI) in healthcare is a field of growing relevance and importance, but in many LMICs, those seeking to develop AI based solutions for healthcare needs, face significant outstanding challenges. This research analysed practical efforts to implement AI-based technologies to support healthcare delivery in low-resource settings. By investigating six pilots within the Foreign Commonwealth and Development Office’s Frontier Technologies program through analysis of associated pilot literature and semi-structured interviews with key pilot actors, we identified differences and commonalities in the experiences of each pilot, and in the perceived enablers and barriers for effective implementation of AI health tools.

We found that AI is a promising tool in this sector but currently lacks the operating environment to be widely successful in solving healthcare challenges. Gaps in regulatory and ethical governance in these contexts exacerbated concerns around the ethical and responsible use of AI and led to alternative technical approaches being followed. The value of partnerships and relationships was demonstrated as essential, and projects with pre-established networks with key decision makers in healthcare systems, both at a bureaucratic and clinical level, demonstrated greater success in both developing and scaling their solutions. The challenge of sustainability and longer-term impact was also identified. The fragmented nature of local technology ecosystems also posed a common barrier to the delivery and scale-up of promising AI tools. It is anticipated that this research can help share some useful lessons for future users and developers of AI technologies and tools in the health space, particularly in resource-constrained settings.

These findings suggest that barriers to equitable AI adoption in low-resource settings are primarily institutional and systemic, rather than technical, highlighting the need for health system–level readiness alongside technological innovation.

**Author Summary:** Artificial intelligence (AI) is increasingly promoted as a way to improve healthcare delivery, including in low- and middle-income countries (LMICs). However, much of the existing discussion focuses on technical performance, with less attention to whether AI tools can be implemented, governed, and sustained within real-world health systems. In this study, we examine a set of AI-for-health pilot projects implemented in low-resource settings to understand what enables or constrains their adoption.

Using interviews with practitioners and a review of project documentation, we explore how these pilots interacted with existing health system conditions, including workforce capacity, data infrastructure, governance arrangements, and institutional partnerships. We find that many of the challenges faced by AI projects are not primarily technical, but instead reflect broader system-level constraints, such as limited regulatory capacity, fragmented data systems, and reliance on external actors for development and maintenance.

Our findings suggest that achieving equitable and inclusive AI for health requires more than developing effective technologies. It also requires sustained investment in the institutions, governance structures, and system capacities that allow AI tools to be safely adopted and integrated into health services. This study offers practical insights for policymakers, funders, and practitioners seeking to use AI in ways that strengthen health systems rather than bypass them.

## Introduction

Artificial Intelligence (AI) has the potential to help solve some of the world’s most pressing challenges, but equitable impact is only possible if the benefits of the technology are relevant, affordable, and accessible to everyone. AI-based tools can help tackle challenges in global public health and have the potential to transform healthcare access and delivery to populations across Low- and Middle-Income Countries (LMICs), but there is considerable concern around the ability to monitor and regulate health AI tools as they are implemented in new contexts. The World Health Organization (WHO) for example has consistently spoken on the role of AI as part of the health ecosystem that can be used to foster safety, equity, and advance the Sustainable Development Goals (1) (2).

A wide variety of tools and solutions have developed, supporting a range of healthcare functions including diagnosis, healthcare planning, monitoring and health communication (3). As healthcare demands grow across countries, and greater volumes of data are available, with the uptake of digital health information systems, the role of tools (both AI/ML powered and otherwise) will become of even more importance and significance to the healthcare sector.

The benefits of using AI in healthcare are potentially significant but need to be weighed against the challenges and risks of implementing solutions. Such benefits include advances in diagnostic accuracy (4), operational efficiency (5), expanded access to care (6), and data-driven population health management (7), illustrate how AI can simultaneously enhance the quality, reach, and sustainability of health systems. However, there remain the presiding risk around of AI solutions exacerbating inequities and perpetuating exclusion. This highlights a need to ensure inherent system biases are not built into systems, including through developing and training systems on inclusive datasets (8).

Across LMIC contexts there can be large differences within populations in terms of access to digital technologies, or digital literacy. These differences risk a ‘digital divide’ where those with existing access benefit from new health innovations, and those without do not. (9) (10). Differences in ‘trust’ of technology can also act as a barrier to uptake (11), although it is worth noting that reservations among populations about trusting the outputs of an AI system over a clinician are often well founded.

The introduction of AI solutions within existing health contexts also risks wider shocks to the system. The concept of “algorithmic accountability” has gained traction when discussing the legal implications and gaps of AI solutions, whereby it is unclear how algorithms or tools are held morally responsible or accountable for decisions regarding a patient’s care, in the same way a healthcare professional can (12). In this regard, the implementation of AI solutions risks disrupting existing accountability mechanisms within health systems. There are also risks that AI solutions lead to the longer term ‘deskilling’ of health workforces, whereby if new solutions replace clinician-led processes, they send signals that there is less demand for skills, which may still be critical for the wider health system. (13)

Whilst there are a wide range of potential issues associated with the implementation of new AI solutions for healthcare use cases, across global contexts, a lack of underpinning regulations, accepted ethical best practice and governance structures for overseeing the training and use of AI based solutions, further inhibits the ability for practitioners to develop and deploy solutions in a safe, responsible and inclusive manner. In LMICs contexts a range of underlying and contextual factors can further exacerbate the potential for these risks emerge. Recent international recommendations have emphasised the importance and need for capacity building and global coordination mechanisms to address these governance gaps (14).

In recent years many early-stage AI solutions have been piloted and trialled in LMIC contexts – with notable challenges around longer-term risks, as well as in relation to the longer-term scale-up and adoption of solutions. This research therefore seeks to understand and answer: *What lesson(s) can be learned from healthcare-focused AI pilots in developing contexts?* Examining a cross-section of pilots and their implementation journeys, challenges, enablers, and wider implications for implementation in health across low-resource settings.

This links also to a widening gap between the Global North and South in terms of access to technologies, systems and skills that will allow these countries to equitably participate in the development and benefit realisation of AI health technologies (15) (16). The presiding literature on this topic demonstrates that much of the research is at a systematic level or applies Western ideas and research to developing contexts and questions. There is therefore a gap in this research landscape for analysis on real-world attempts to utilise and integrate AI technologies in healthcare systems within developing contexts. Closing this research gap will be imperative to support future innovators to develop their own plans or pilots for similar schemes as well as begin to build a research base for practical implementation of AI in these contexts that is currently not present.

Research analysing activities using AI in resource-poor settings is aligning to the idea that AI should look to build intelligence into existing systems and institutions rather than starting from scratch or hoping to replace existing systems, however broken these may be (17) (18). An AI system is dependent on these underlying systems being available with data it can learn from, but if these existing systems are insufficient, effective will be inhibited and the ethical deployment of AI tools in these contexts may be questioned. There is therefore a need to ensure systems are appropriate for AI implementation. For an AI application to be successful, activities require knowledge of local markets, clear usability requirements and access to adequate training data via field testing (19) (20). However, despite the presiding gaps in robust evaluation and impact measurement of interventions, there is a growing need to supplement this evidence base and provide insight into how AI is currently being tested and applied and their potential to be integrated into supporting systems.

This research aims to support the filling of this literature gap by analysing practical attempts to implement AI based technologies for health outcomes in low-resource or developing settings, to support policymakers and practitioners in the design and development of their own programs using AI to help them consider the identified enablers and challenges of doing such projects.

### Research aims and Rationale

This research aims to look at 6 pilots that explored the application of AI innovations for health use cases in developing contexts to identify differences and commonalities in their design and delivery and what lessons could be gleaned for wider adoption and implementation of AI for health tools in comparable contexts. This research analysed six pilots supported and funded by the Frontier Technologies Hub (FT Hub), that utilised AI for health in a range of developing contexts, to identify any differences or commonalities in their design and delivery and what lessons could be gleaned for wider adoption and implementation of AI for health tools in comparable contexts.

Beyond documenting implementation experiences, this study makes a conceptual contribution to debates on global inequities in AI-enabled health systems. Drawing on an institutional perspective, it shows that barriers to inclusive AI adoption are not primarily technical, but reflect deeper asymmetries in regulatory capacity, data infrastructures, and governance between global innovation models and LMIC health systems. By analysing how AI pilots navigate these institutional conditions, the paper advances digital health implementation research by demonstrating that equity-oriented AI adoption is fundamentally a systems challenge rather than a problem of technological performance.

### About the Frontier Technologies Hub

The Frontier Technologies Hub works with UK Foreign, Commonwealth and Development Office (FCDO) to understand the potential for innovative tech in the development context and then test and scale the ideas. It invests in early-stage ideas and radical technology solutions, build capacity to develop, use and govern technology responsibly and support locally led innovation.

## Results and Discussion

This research has revealed that for AI solutions to be implemented effectively, a *whole systems* approach is needed—one that looks across multiple dimensions of the health ecosystem and addresses the specific needs within each. Such an approach increases the likelihood that interventions can scale in ways that are impactful, inclusive, and responsible. Participants’ reflections were grouped under the following themes:

- The importance of taking a user-centred approach
- Building effective partnerships
- Establishing a wider operational model around a solution
- The value of an enabling policy environment
- The wider technology and data environment

Whilst these themes reflect the experiences of individual pilots, they also point to broader systemic conditions shaping AI-for-health implementation in LMICs. Interpreted through an implementation science lens, the findings map closely onto frameworks such as the WHO Health System Building Blocks, highlighting how governance, workforce capacity, data systems, and organizational readiness interact to enable or constrain adoption.

Collectively, the findings from this research extend beyond the individual project or case-study experiences, to illustrate how AI and health initiatives are shaped by structural and institutional conditions within LMIC health systems. The recurring challenges observed across the pilots investigated point to systemic inequities in data readiness, regulatory governance, and institutional capacity, highlighting that successful AI adoption depends as much on enabling environments as on technical innovation.

[Pilots have been assigned an anonymous moniker throughout this section – for example Pilot (P)1.]

### The importance of taking a user-centred approach

The pilots analysed identified the value of user research in helping them develop solutions which were responsive to user needs and the problems experienced within health systems. For the pilot P1, the pioneers undertook a user centred approach by engaging healthcare actors early on to support its transition from an assumed problem definition to a completely new focus – whereby it shifted towards developing a solution that could help healthcare workers to identify where vulnerable individuals had stopped accessing services, rather than trying to identify ‘the most vulnerable’ amongst those already accessing services.

For pilot P2, research participants explained how not exploring user research considerations early enough – including looking to comprehensively understand the root causes behind the problems being faced by patients - hindered their ability to deliver a service effectively. The research participants discussed how they believed earlier research to understand norms and beliefs of end users would have really helped to inform the solution. Such user-centred approaches have been shown to improve uptake and usability in LMIC digital health deployments (21). Pilots across the study surfaced challenges adopting a user-centred approach, including around identifying existing technical tools – especially LLMs with indigenous language support – that could be reasonably applied and adapted for local user needs.

- *“The language learning models in [the local language] were fine, but for the indigenous languages, it didn’t work.”*

More generally, those pilots that involved early user research and iteration, achieved better engagement outcomes, and encountered fewer problems as the pilot progressed:

- *“If you want AI to succeed in LMICs, you have to start with the user — from language to literacy to how they actually access healthcare.”*

P7 for example, was a tool explicitly designed to address access barriers faced by rural populations, as well as targeting users with limited health knowledge or resilience, reinforcing equity considerations. This emphasis on lived experience reinforces the view that relevance and uptake depend on aligning AI tools with users’ everyday constraints, rather than optimising technical functionality alone.

- *“People who live in the mountains have to spend around days on a bus to go into the hospital in [city] for a 10-minute appointment (…) our technology was designed to help people who don’t have that type of resilience.”*

From a systems perspective, these findings illustrate that user-centred design is a proxy for deeper alignment with service delivery realities and workforce capabilities. In the absence of meaningful engagement with frontline health workers, AI tools risk misalignment with existing workflows and capacity constraints, limiting adoption regardless of technical sophistication. This reflects broader implementation challenges observed in digital health, where technologies fail not due to lack of innovation but due to insufficient integration with human and organisational systems.

### Building effective partnerships to enable action and knowledge sharing

Local partnerships and co-designing solutions are frequently discussed in academic literature as being critical enablers of effective digital health implementation in LMIC settings (22) (23). Almost all pilots highlighted the value of partnerships—particularly in establishing trust and collaboration between innovators, healthcare providers, and governing bodies—as a prerequisite for developing, testing, and scaling solutions. For P1, early engagement with the government health body was critical for:

- Access to health professionals, with whom they could engage to understand user needs and priorities and test the innovation.
- Access to relevant data for training and developing AI solutions.
- Receiving critical feedback, and guiding development decisions, to ensure the solution would help tackle priorities within the region, and the issues that cause the greatest amount of harm.
- Securing formal adoption of new solutions amongst health staff, including within everyday processes and workflows – in this case the governing body made formal issuances for the use of the tool.

In cases where pilots failed to secure partnerships with key system actors, their ability to integrate and test solutions was severely limited. For example, P3 attempted to work with both their country’s’ Ministry of Health and hybrid public–private hospitals, but neither organisation had the processes or risk appetite to onboard a pilot. As a result, the team could not test their solution with the most vulnerable groups—such as rural and low-income populations—who faced the greatest barriers to healthcare and were the intended beneficiaries. Risk aversion and a perceived lack of trust in AI technologies, and in the ability to implement solutions in an ethically compliant way, proved to be a barrier to developing partnerships for this pilot – although the pilot showed that through proper implementation these concerns can be addressed. Partnerships between domain and technical experts also played a critical enabling role in many of the pilots: P4 for example, brought in subject matter expertise, whilst P5 cited a need for greater clinical involvement in development of solutions. Other types of partnerships were seen as critical, particularly the importance of partnerships between peers, whereby people working on AI solutions across different LMIC contexts could share learning with one another. P5 emphasised the importance of peer learning between African start-ups, but not with Silicon Valley. For P3, an underlying driver behind the inability to form a partnership appeared to be lack of trust and risk aversion – with health provider organisations having scepticism that the pilot team would be able to build and test technology in a lean way whilst still being ethically compliant. Finally, P7 noted how its partnerships were said to be essential for navigating institutional complexity, particularly where local regulatory pathways or implementation authority were fragmented.

These findings suggest that partnerships function not only as operational enablers but as mechanisms for navigating institutional authority and legitimacy within health systems. Access to data, ethical approval, and formal adoption was mediated through state and professional actors, underscoring how power is distributed within health governance structures. For externally funded or technologically driven initiatives, this highlights how North–South asymmetries in expertise and resources must be negotiated through locally grounded partnerships to avoid exclusion from core health system processes.

### The importance of establishing the operational model, and ensuring health workforce needs for adopting the solution are met

AI technologies have limited impact unless adopted as part of a process which considers how solutions get integrated within existing operational models. This includes working to understand how the solution might work alongside and within existing operational processes or even transform those processes.

Solutions that were effectively implemented looked beyond the technology, to identify what changes were required in this space. In the case of P1, this involved not only integrating use of the solution within existing health actor workflows but actually using the technology to enable the implementation of new workflows and processes that previously were not possible. In particular, their solution through making it possible to produce a detailed list of vulnerable individuals in the community requiring direct support to access healthcare, they could introduce new processes where for public health aides (community health workers) directly engaged these individuals and helped them to ‘navigate’ and access basic services (24).

Strong partnerships were strongly linked to project success, but misalignment in expectations was shown to slow progress. Resistance from government institutions or funders due to political factors and risk aversion was identified as a barrier, with some projects experienced tension between local organisations and global donors due to differing priorities:

- *“Public institutions are very risk-averse… there is no such thing as an investment environment for pilots.”*
- *“These solutions can’t be designed on their own - ecosystems really matter.”*

A critical consideration identified by pilots which engaged directly with health workforce actors, was the need to consider the skill level of these actors to implement the technology, or their needs for skill development where necessary. P1_ found that despite the implementation of a digital platform it’s adoption among many health actors, but especially community health workers were limited, and that these actors lacked awareness or capacity to use the technology. Consequently, training was provided on the use of the technology within their everyday processes.

As well as providing support to healthcare workers to develop the skills to use technology in their working roles, one pilot identified the value in upskilling healthcare workers so that they could play a critical role in the research and development of solutions. Likewise, P2 involved clinicians in the testing and evaluation of their solution and identified a need to upskill a subset of the health workforce so that they could play a more central and active role in the training and verification of AI solutions. P7 similarly developed operational models that complemented existing clinical workflows, rather than replacing them, were perceived as more acceptable by implementers, positioning AI as a support mechanism for the health workforce, rather than as a substitute for clinical expertise:

- *“We created digital selves for patients and the clinician… to automate routine services.”*

Many pilots grappled with defining where AI could automate tasks effectively and where human judgment remained essential. In the case of P6 using an AI solution for diagnostics, the solution was effective at diagnosing certain conditions but not others, highlighting the need for ongoing human oversight. Comparatively, P4 and P5 acknowledged the limitations and risks of AI solutions, in place of existing human led processes, and decided to scope their solutions so that it was not undertaking activities previously undertaken by a trained clinician. Across all examples, successful approaches built operational models where AI efficiency was paired with expert review, supported by processes for continuous monitoring of outputs.

These challenges reflect tensions at the organisational and wider system levels, where AI adoption interacts with professional roles, accountability structures, and long-term sustainability. Concerns around deskilling and accountability highlight that AI interventions are not neutral additions but reshape labour relations and institutional responsibilities. Without parallel investment in workforce capacity and governance mechanisms, AI risks destabilising rather than strengthening health systems.

### The value of an enabling regulatory environment

A critical need identified by several pilots was the need and value of guiding regulations to help inform the research, design and testing of new interventions. With multiple pilots citing how they felt they benefited from being able to adopt existing guidelines, and others citing how an absence of guidelines and rules inhibited their ability to make progress and feel confident they were delivering solutions in a responsible way.

Two pilot team P1 & P3 identified that following existing Institutional Review Board (IRB) processes was valuable as it set clear rules on how the team needed to approach the research and development of the technology and enabled them to test their solutions in ways which mitigated potential for harm. Comparatively, P2 didn’t plan to implement ethical guidelines until later in their work, and some team members cite this as something that inhibited progress early on. P7 circumvented this challenge by acquiring ethics approval via a high-ranking university in country, but it was noted that engagement with the local and national public health institutions here was more vulnerable to political instability and hierarchical decision-making structures:

- *“Even if they ran the local hospital, they didn’t have the power to make decisions because they always had to refer up.”*

In addition to regulations to guide best practice research and testing of innovations, a critical factor barrier identified by teams to implementing AI solutions in a responsible and ethical manner was the lack of regulation relating to the specific risks of AI solutions – including issues around inaccuracy, bias, accountability, data privacy, safety, and misinformation.

This led to challenges on pilots, including how to effectively validate accuracy, before determining whether it was safe to trial the AI solution with patients.

- *“We couldn’t agree on how to check AI accuracy—whether it should be reviewed by five medical professionals or have ongoing audits.”*

In the case of P4, a decision was made to adapt the proposed solution, and remove generative AI component, due to concerns of solution inaccuracies, hallucinations and biases that might result in longer term harm.

P6, identified risks around ‘accountability’, where an AI solution was replacing human processes, where individuals were clearly accountable for performing a clinical function, with an AI solution, where it was unclear who would be accountable, in the event it made inaccurate predictions. The pilot also identified risks that their solution might lead to ‘deskilling’ of the existing workforce, and/or send signals that there is less demand for skills - and therefore destabilising the system in which there being implemented. Critically the pilot acknowledged the challenge of effectively moving forward with a proposed innovation, without regulation in place to help navigate these types of complex ethical issues.

For many pilots it was clear that AI innovation had outpaced regulation, with P5 looking to European Union (EU) AI regulations, to identify how their solution might be integrated into public health systems, while identifying the limitation that international guidance often lacks contextual relevance. This resonates with academic discourse that argues argue for regional approaches to digital sovereignty that better reflects African contexts, rather than transplanted international rules (25) (26).

Within some of the Frontier Technologies pilots, the disconnect between local regulatory expectations and international best practices, led to project delays. The lack of formal AI guidelines in some LMICs meant teams had to define their own ethical frameworks, often with differing perspectives across teams and with the funding organisation.

- *“We really didn’t align on the standards and safeguards that would need to be in place to monitor and test the accuracy [of the solution].”*
- *“Obviously, because it’s all AI and it was new, there was no such thing as regulation yet.”*
- *“There’s a balance between adding value and managing risk—if it’s too restrictive, it’s not useful, but if it’s too open, it could be dangerous.”*

Several pilots highlighted a regulatory gap in African contexts, calling for an African Union–level framework similar to EU AI regulations. They stressed that while such frameworks could promote safer and more accountable innovation, the costs of meeting responsible AI standards are high and often underestimated. The absence of contextually appropriate AI regulation effectively shifted ethical responsibility from institutions to individual implementers, requiring pilot teams to develop ad hoc governance mechanisms. This reflects broader debates on digital sovereignty in LMICs, where global regulatory models often fail to account for local capacities and priorities. As a result, responsibility for managing AI risks becomes unevenly distributed, reinforcing global inequities in who bears the burden of ‘responsible’ innovation.

### The value of an enabling technology and data environment

Participants found that to develop AI solutions, there was a requirement to access good quality and representative data. Where this was not possible it inhibited the delivery of more comprehensive AI solutions that could deliver benefits such as predictive modelling / diagnostics. This challenge was faced by P1 which encountered issues around both duplication of records, but also misaligned data sets that were not interoperable and could not be combined to create a single data set for AI analysis. The pilot identified the need for foundational work, to establish data sharing and stewardship practices, and enable interoperability between disparate and siloed datasets.

Whilst a lack of interoperability was a problem for certain pilots, one pilot (P3) working in an environment with an absence of legacy digital systems or entrenched technologies for generating, processing and storing digital data, identified that this created an opportunity rather than a challenge. In this regard, an absence of contractual and technical blockers to data integration, proved an enabling factor.

Data constraints observed across pilots reflect not only technical limitations but structural patterns in global data production and ownership. Fragmented, non-interoperable datasets limit local capacity to develop and govern AI systems, while reliance on externally developed platforms risks reinforcing forms of data dependency. These dynamics echo concerns around data colonialism, where LMICs contribute data but retain limited control over its downstream value and governance. These dynamics echo concerns around ‘data colonialism’, where LMICs contribute data but retain limited control over its downstream value and governance (27).

### The need for a systems approach

Scaling digital health in LMICs requires attention to ecosystems and sustainable business models, not just pilot efficacy (28). The findings across each of the pilots indicate that a broad range of considerations are required to effectively develop and test early-stage AI innovations for health. Approaches which look beyond technical implementation, to a breadth of additional changes – including changes to partnerships, to operational processes, to capacity building, to wider regulations – necessary for technology to be scaled in an effective, sustainable and responsible way. Although all pilots analysed for this study were at an early stage in the innovation process, the adoption of a broader approach proved essential for facilitating the delivery of new solutions and enabling their potential for sustainable scale-up.

Taken together, the recurring barriers observed across pilots are best understood as manifestations of systemic conditions rather than project-level failures. Weak regulatory capacity, fragmented data infrastructures, constrained workforce readiness, and asymmetrical global innovation relationships collectively shape the feasibility of AI-for-health implementation in LMICs. These findings suggest that inclusive AI adoption depends less on individual pilot design and more on investment in institutional readiness, local governance, and digital sovereignty. Recurring implementation barriers were not isolated to individual projects but reflected broader system-level conditions related to governance, infrastructure, partnerships, and workforce capacity. This was exemplified by P7, who spoke about the arrest of a senior political figure in their country of practice, and how the downstream impact of this led to a pause in activities for six to eight months. This underscores the importance of viewing AI-for-health pilots as embedded within dynamic political and institutional systems, rather than as stand-alone technological interventions.

## Conclusion

This research has unearthed the experience and thoughts of key stakeholders who have sought to pilot and develop AI tools to support healthcare delivery in LMICs. It has sought to set out the key experiences of these pilots and its contributors, in the hope that these can guide future pioneers of AI technologies and tools in the health space. It became clear through this research, that AI is promising but it is not a magic bullet. Several pilots began with a plan to deliver AI solutions with autonomy to adopt specific health system functions, but in the end opted for less autonomous, and more managed solutions, because of technical and ethical concerns around AI. This was attributed to factors including a lack of regulatory or ethical governance for AI use in their countries, as well a lack of supporting data infrastructure, to enable more seamless adoption of AI. AI-driven tools will continue to be challenged by a lack of clear regulatory frameworks in LMICs, and Governments must seek to provide that certainty as an imperative, if they are to develop bountiful AI ecosystems in their healthcare sectors.

Across the pilots, the value of partnerships and relationships proved a key lever for success when implementing, integrating and scaling new tools into pre-existing health systems. Integration of key actors, particularly decision makers in relevant organisations, was critical for access to patients and data as well as ethical approval, which was a key blocker for pilots without these networks. Also, incorporation of healthcare professionals at the earliest stages were a key factor for success, to understand user needs and to receive critical feedback, to ensure the core aim of a pilot or tool was actually meeting healthcare needs. Both of these groups of actors are also imperative for scaling and integration into everyday processes and workflows in the longer term.

Finally, sustainability and long-term impact measurement were also identified as weak points across the pilots, with the majority of pilots lacking clear pathways for scaling or tracking real-world health outcomes. It was clear that many of the pilots struggled to identify methods of assessing AI efficacy at an early stage that could be used to build a case for its further piloting. This is partly because pilots lacked an ability to track metrics, or an ability to attribute identified changes to the pilot intervention.

Future research should seek to explore further the issues around accessibility and bias in developing tools and products and ask, how might we enhance the scalability of AI technologies in diverse socio-cultural contexts. There is also a need to understand how we ensure that the benefits of these technologies are accessible to the most marginalised communities. Answers to these will undoubtably unearth further questions and challenges but will contribute unquestionably to the research landscape for what is sure to be a burgeoning topic for Governments and technology pioneers in the coming decades.

## Materials and Methods

This research selected the programs shown in [**Table 1**] for investigation within this study. These pilots and programs were selected as part of the Frontier Technologies Hub portfolio, which tests novel and innovative technologies to solve development challenges. The aim here is to answer the key research question:

- What were the key challenges and enablers in these programs when implementing AI-based innovations within health systems in developing contexts?
- And from this, what lessons can be learned for future AI for health-based programs in developing contexts?

**Table 1:** Pilots included in the study.

| Pilot Name | Country/Region pilot delivered in | Pilot still ongoing when research was conducted? |
| --- | --- | --- |
| eTriage for social inclusion: navigating out of vulnerability situations | Philippines | Yes |
| EmpatIA - AI to enhance healthcare in remote areas of Peru | Peru | Yes |
| AI-assisted tools to Diagnose TB and Silicosis | South Africa | No |
| mDoc (COVIDaction) | Nigeria | No |
| Developing a chatbot SMS/IM for health behaviour change in Africa | Nigeria | Yes |
| Sexual Reproductive health chatbot | Kenya | No |

Initial desk-based research was conducted to analyse the current literature that investigates trends in applications of AI for health (including role and use cases), programs or projects that use AI as a core function or focus and common use cases explored for AI in health in developing contexts (LMIC/LDC focus), examples where AI has been scaled for impact in global health systems in LMICs, and any observed challenges and enablers for effective piloting and scaling of health innovations in LMICs – including as they relate to different building blocks within health systems (as defined by WHO building block framework). This was used to inform a literature review and develop and understanding on the current predominant research on AI for development and health and to guide the development of the research questions and semi-structured interviews.

## Qualitative Methodology and Sampling

Seven semi-structured key informant interviews (KIIs) were conducted with pre-identified individuals who contributed to the selected projects that utilised AI as a technology to support a healthcare goal in a low-resource setting. These interviews sought to identify differences and/or commonalities in the experience of AI health pilots, and what lessons could be gleaned for wider adoption and implementation of AI for health tools in LMIC contexts. These KIIs were conducted with respondents who occupied one of the following categories:

- Technologists involved in developing and testing AI-based digital tools in the pilot/s.
- Coaches who helped to facilitate pilots without taking responsibility for direct implementation

To note, that data and discussions in the following sections from the KIIs are not attributable to the pilot the participant worked on. This anonymity allowed for deeper data collection with participants feeling comfortable to be open and honest particularly around the challenges their pilot/s faced.

The sampling criteria and recruitment rationale was based upon pre-existing networks of colleagues that had direct experience of designing, implementing, or supporting AI pilots to achieve healthcare outcomes or goals in LMIC contexts. Eligible participants were required to have been actively involved in one or more of the identified Frontier Technologies Hub pilots, either as technical developers, programme implementers, or delivery coaches providing oversight and support. Participants were identified through project documentation and programme records and were invited via email to participate in a semi-structured interview. This approach was selected to ensure inclusion of perspectives across technical, operational, and governance dimensions of AI implementation. The sample size is consistent with qualitative implementation studies seeking depth of insight rather than representativeness. Saturation was assessed iteratively during analysis, as recurring patterns related to partnerships, governance, data availability, and operational integration were consistently identified across participants.

### Data Collection and analysis

The authors initially conducted a desk review of existing literature that relates to the study questions, including reports and blog posts from the Frontier Technologies program pilots analysed in this research.

Following this, the authors conducted the KIIs with participants and contributors to the pilots and programs. These KIIs covered the extent to which existing solutions have enabled usage and engagement by users and patients with developed tools and products, as well as lessons from technical implementation on enablers and barriers. Content analysis was conducted using AtlasTI© to identify themes and trends in responses. An initial coding framework was developed deductively from the literature review and research questions and inductively refined through close reading of transcripts. Coding was conducted by at least two of the listed authors per KII transcript to validate coding consistency and interpretation. Themes were generated through iterative comparison across interviews and triangulated with findings from the desk review and pilot documentation. Reflexivity was addressed through regular analytic discussions among the authors, acknowledging the research team’s prior engagement with the programme context.

## Data Availability

Data including codebook and anonymised transcripts are available upon reasonable request.

## Acknowledgments

The authors want to thank all the participants across the different pilots who contributed their time and insights to this work. Thank you also to colleagues across the Frontier Tech Hub, DT Global, and FCDO for their contributions and guidance.

## Author Contributions

CM and IV developed the core study including the proposal and methods. CM and JM supported the ethics process alongside the funding agreement which IV also supported. CM collected the data for the KIIs. CM and IV both conducted the thematic analysis on the data and collectively agreed the results and key findings. All authors had a role in writing the manuscript through to its final version.

## Ethics

Ethical approval was granted by the University College London Research Ethics Committee (Approval ID: 28293/002) and covered all interviews conducted with participants, regardless of their geographic location, as data collection was undertaken remotely and did not involve direct engagement with patients or communities in the pilot countries.

## Funding Statement

Research funded by DT GLOBAL INTERNATIONAL DEVELOPMENT UK LTD. T&Cs set out in grant reference no. 21108 – Frontier Technologies 2.

## Notes

### Competing Interest Statement

The author IV previously worked for Results for Development, an organisation affiliated with DT Global, which manages the Frontier Technologies Hub programme referenced in this study. IV was not involved in the funding, commissioning, or management of the research pilots reported in this article. All other authors declare no competing interests.

### Funding Statement

Yes

### Author Declarations

University College London Research Ethics Committee (Approval ID: 28293/002)

